# Comparisons of the rate of acute myocardial infarction between COVID-19 patients and individuals received COVID-19 vaccines: a population-based study

**DOI:** 10.1101/2022.07.25.22277985

**Authors:** Oscar Hou In Chou, Cheuk To Chung, Danish Iltaf Satti, Jiandong Zhou, Teddy Tai Loy Lee, Abraham Ka Chung Wai, Tong Liu, Sharen Lee, Vassilios S Vassiliou, Bernard Man Yung Cheung, Gary Tse

## Abstract

**Background:** Both Coronavirus Disease-2019 (COVID-19) infection and COVID-19 vaccination have been associated with the development of acute myocardial infarction (AMI). This study compared the rates of AMI after COVID-19 infection and among the COVID-19 vaccinated populations in Hong Kong.

**Methods:** This was a population-based cohort study from Hong Kong, China. Patients with positive real time-polymerase chain reaction (RT-PCR) test for COVID-19 between January 1^st^, 2020 and June 30^th^, 2021 were included. The data of the vaccinated and unvaccinated population was obtained from the “Reference Data of Adverse Events in Public Hospitals” published by the local government. The individuals who were vaccinated with COVID-19 vaccination prior the observed period (December 6^th^, 2021 to January 2nd, 2022) in Hong Kong were also included. The vaccination data of other countries were obtained by searching PubMed using the terms [“COVID-19 vaccine” AND “Myocardial infarction”] from its inception to February 1^st^, 2022. The main exposures were COVID-19 test positivity or previous COVID-19 vaccination. The primary outcome was the development of AMI within 28 days observed period.

**Results:** This study included 11441 COVID-19 patients, of whom 25 suffered from AMI within 28 days of exposure (rate per million: 2185; 95% confidence interval [CI]: 1481-3224). The rates of AMI were much higher than those who were not vaccinated by the COVID-19 vaccine before December 6^th^, 2021 (rate per million: 162; 95% CI: 147-162) with a rate ratio of 13.5 (95% CI: 9.01-20.2). Meanwhile, the rate of AMI was lower amongst the vaccinated population (rate per million: 47; 95% CI: 41.3-53.5) than COVID-19 infection with a rate ratio of 0.02 (0.01, 0.03). Regarding post-vaccination AMI, COVID-19 infection was associated with a significantly higher rate of AMI than post-COVID-19 vaccination AMI in other countries.

**Conclusions:** COVID-19 infection was associated with a higher rate of AMI than the vaccinated general population, and those immediately after COVID-19 vaccination.

## Introduction

Since 2019, the emergence of severe acute respiratory syndrome coronavirus 2 (SARS-COV-2) has led to increased mortality, socioeconomic disruption, and overloading of the healthcare sector [1-3]. The exponential rate of disease spread has prompted the need to develop risk stratification strategies, treatment and prevention efforts against the Coronavirus Disease-2019 (COVID-19) [4, 5]. One of these efforts was achieving universal access to COVID-19 vaccines. Recent studies have shown promising effects of various vaccines, including reducing the severity of symptoms and boosting individual immunity [6, 7].

Despite the large-scale success of compliance in some countries, monitoring vaccinated individuals reveals the occurrences of relatively rare but significant complications, including anaphylaxis, Ramsay Hunt syndrome, cerebral venous thrombosis, and cardiovascular thrombotic events [8, 9]. Acute myocardial infarction (AMI) was one of the thrombotic events that emerged amongst COVID-19 vaccine recipients, especially in older populations. Numerous studies, including randomised controlled trials and cohort studies, have reported on the occurrence of AMI [10-14]. A pooled analysis demonstrated that the majority of AMI patients post-vaccination developed symptoms after the first dose of the BNT162b2 vaccine [15]. Similarly, a systemic review study found that thromboembolic events were commonly reported in patients who were administered with the AstraZeneca vaccine, with cerebral venous thrombosis being the most prevalent manifestation [16]. Nevertheless, it is still too early to draw a definitive causal relationship between the COVID-19 vaccine and AMI. Hence, further investigation is necessary to understand the nationwide trends of AMI fully.

Indeed, there has been limited exploration of the temporal association and differences in AMI risk between infected cases and vaccinated individuals. By understanding potential severe complications, this will allow us to improve upon current vaccination interventions and ultimately minimise the likelihood of cardiac manifestations. Thus, the aim of the present study is to compare the risk of AMI between COVID-19 patients and vaccinated individuals.

## Methods

This population-based retrospective cohort study was approved by the Institutional Review Board of the University of Hong Kong/Hospital Authority Hong Kong West Cluster (UW 20-250). The need for informed consent was waived by the Ethics Committee owing to its observational and retrospective nature. Patients who tested positive for COVID-19 by real-time polymerase chain reaction (RT-PCR) at any of the Hong Kong public hospitals or outpatient clinics between January 1^st^, 2020 and June 30^th^, 2021 regardless of their vaccination status were included. The data were obtained from the local electronic healthcare database, Clinical Data Analysis and Reporting System (CDARS), as reported previously [17-19]. The primary outcome was AMI within 28 days of COVID-19 disease. The patients’ demographics and prior comorbidities were also retrieved.

The vaccination data of other countries were extracted using keywords labelled “The literature search was done in PubMed and Embase by keywords “COVID-19 vaccine” AND “Myocardial infarction” until February 1^st^, 2022. A total of 15 articles were identified by this search term.” upon searching PubMed, Embase and the official reports of Hong Kong. Papers that did not contain the information regarding COVID-19 vaccination or AMI were excluded. Conference papers or abstracts, reviews, systematic reviews, meta-analysis were also excluded. The rate of AMI in the vaccinated individuals and unvaccinated individuals in Hong Kong within 28 days of exposure were extracted from the “Reference Data of Adverse Events in Public Hospitals” published by the Hong Kong government was up to January 2^nd^, 2022. The COVID-19 vaccinated population in Hong Kong was defined as the population with 1st dose of COVID-19 vaccine since the beginning of the COVID-19 vaccination programme before the observed period (December 6th, 2021). The non-COVID-19 vaccinated population was defined as the individuals as the population without any dose of COVID-19 vaccine since the beginning of the COVID-19 vaccination programme before the observed period. The rates of AMI after COVID-19 infection were calculated by dividing the number of patients by the number of RT-PCR positive COVID-19 patients. The rates of AMI amongst the vaccinated and unvaccinated population were calculated by dividing the cases to the number of the susceptible population. The hybrid Wilson/Brown method was used to calculate 95% confidence intervals (CI) for rates. The rate ratio was calculated to compare the rates of AMI across different groups. In addition, a sensitivity analysis was performed to verify the rate ratio of AMI events after COVID-19 infection and vaccination compared to the background rate across different years. The analysis was conducted using PRISM (Version: 9.0.0) and RStudio (Version: 1.4.1103).

## Results

Among the 11441 COVID-19 patients included in our cohort from Hong Kong, 25 patients developed AMI within 28 days after COVID-19 infection (rate per million: 2185; 95% CI: 1481-3224; **Table 1**). Compared to the individuals who were not vaccinated prior December 6^th^, 2021, the rate was significantly higher with a rate ratio of 13.5 (95% CI: 9.01, 20.2). The clinical characteristics of the AMI patients are presented in **Table 2**. The mean age of the patients was 69.11 years old (standard deviation [SD]: 14.85) and 72% of the patients were male. Meanwhile, among the 4914894 individuals who were vaccinated between December 6^th^, 2021 and January 2^nd^, 2022, 231 patients developed AMI within 28 days after the date of vaccination (rate per million: 47.0, 95% CI: 41.3, 53.5). Moreover, the rate of AMI was a lot higher in COVID-19 patients than in those who were vaccinated (rate ratio: 46.5; 95% CI: 30.8, 70.2).

**Table 1.** Incidence rate and rate ratio of acute myocardial infarction after COVID-19 infection and among the vaccinated population.

| Outcomes | Non COVID-19 vaccinated population (Hong Kong, China)[58] | Covid-19 infection (Hong Kong, China) | COVID-19 vaccinated population (Hong Kong, China)[58] |
| --- | --- | --- | --- |
| Type of vaccines | - | - | CoronaVac<br>BNT162b2 |
| Cases | 405 | 25 | 231 |
| Persons | 2500000 | 11441 | 4914894 |
| Follow-up period | 28 days | 28 days | 28 days |
| Rate per million persons or doses per 28 days (95% confidence interval) | 162<br>(147, 179) | 2185<br>(1481, 3224) | 47.0<br>(41.3, 53.5) |
| Rate ratio (95% confidence interval) | 1 | 13.5<br>(9.01, 20.2) | 0.290<br>(0.247, 0.341) |

**Table 2.** Baseline characteristics for COVID patients that had AMI within 28 days.

| Demographic variables | Mean ± SD / Number and Percentage (n = 25) | |
| --- | --- | --- |
| Age | 69.11 ± 14.85 | |
| Male | 18 | 72 |
| **Comorbidities** |  |  |
| Hyperlipidaemia | 0 | 0 |
| Gastrointestinal bleeding | 3 | 12 |
| Acute myocardial infarction | 2 | 8 |
| VT/VF/SCD | 0 | 0 |
| Cognitive dysfunction | 1 | 4 |
| Anxiety disorder and depression | 0 | 0 |
| Ischemic heart disease | 7 | 28 |
| Sleep disorders | 1 | 4 |
| Hip fractures | 1 | 4 |
| Accident fall | 6 | 24 |
| Alcoholism | 0 | 0 |
| Tobacco use and dependence | 0 | 0 |
| Obesity | 1 | 4 |

In Israel, the rate of AMI after COVID-19 infection was significantly higher than that within 42 days immediately after COVID-19 vaccination [20] **(Figure 1; Supplementary Table S1)**. The rates of AMI amongst elderly patients in different countries were compared. When compared to the COVID-19 patients, the rate of AMI was significantly lower among the elderly ≥70 years old in Israel [20] (rate ratio: 0.097; 95% CI: 0.056, 0.169) and those ≥75 years old patients in France [21] (rate ratio: 0.240; 95% CI: 0.162, 0.357) **(Figure 2; Supplementary Table S2)**. Overall, in Hong Kong, the absolute number of people vaccinated and who were younger than 70 years old, was higher than the absolute number of those more than 70 years old who were vaccinated. As such, COVID-19 patients ≤70 years old were matched with the relatively younger vaccinated population. There was a total of 10276 COVID-19 patients who were below 70 years old (mean age: 40.93; SD: 16.97), 48% of whom are male. Among those patients, 13 COVID-19 patients younger than 70 years old developed AMI (rate per million: 1265; 95% CI: 749, 2163). The baseline characteristics of the 13 patients (mean age: 57.08; SD: 16.56) are presented in **Supplementary Table S3**. Amongst the 13 AMI patients, one patient had a history of AMI, and 2 patients had history of ischaemic heart diseases. The rate of AMI was higher among the COVID-19 patients below 70 years old than among the vaccinated population (rate ratio: 26.9; 95% CI: 15.4, 47.0; **Figure 3; Supplementary Table S4**). The sensitivity testing revealed the same trend that could be observed while comparing the rate of AMI with the whole population across the COVID-19 and pre-COVID-19 era **(Figure 4; Supplementary Table S5)**.

**Figure 1.**
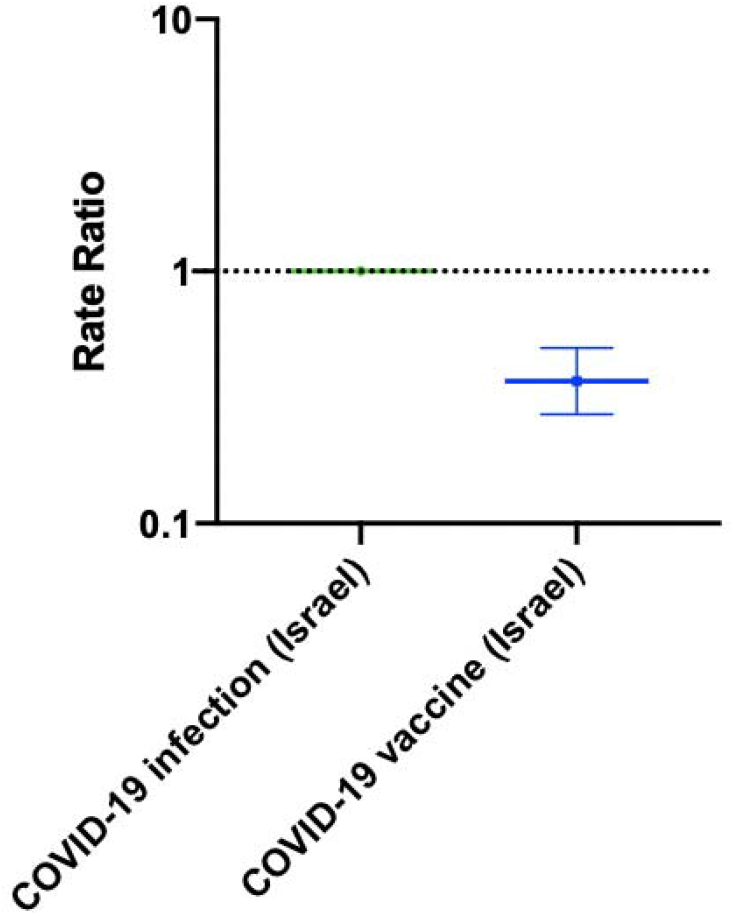
The rate ratio of the acute myocardial infarction events after COVID-19 infection and COVID-19 vaccination within 42 days in Israel.

**Figure 2.**
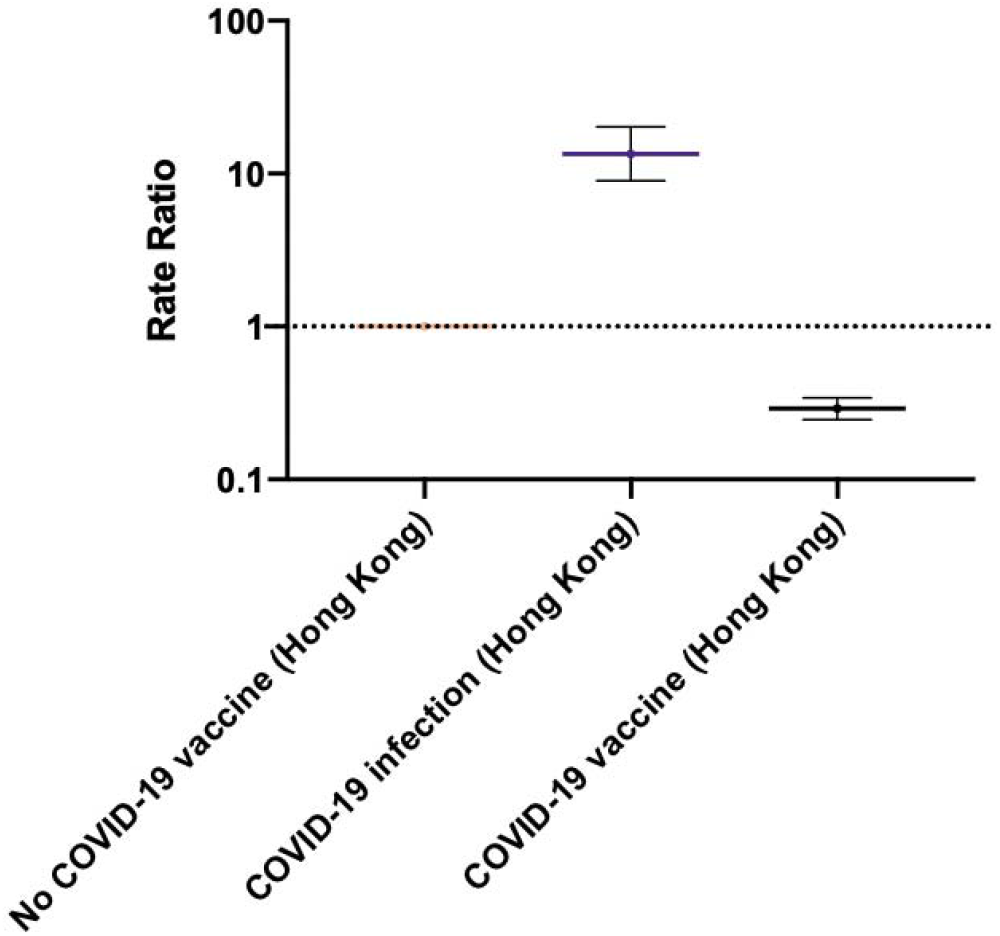
The rate ratio of the acute myocardial infarction events after COVID-19 infection and COVID-19 vaccination in Hong Kong compared to without COVID-19 vaccination.

**Figure 3.**
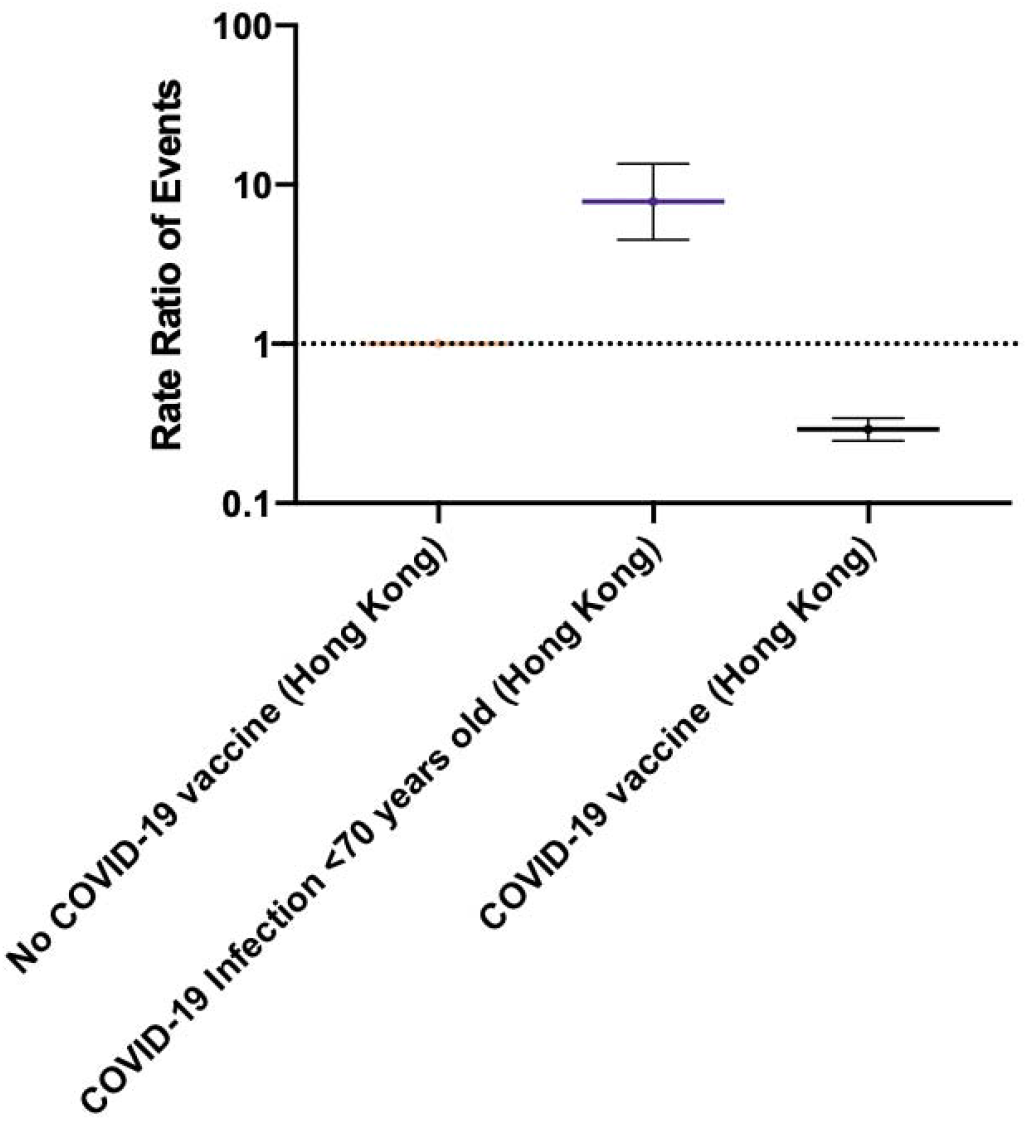
Subgroup analysis: The rate ratio of the acute myocardial infarction after COVID-19 infection under 70 years old and COVID-19 vaccination in Hong Kong.

**Figure 4.**
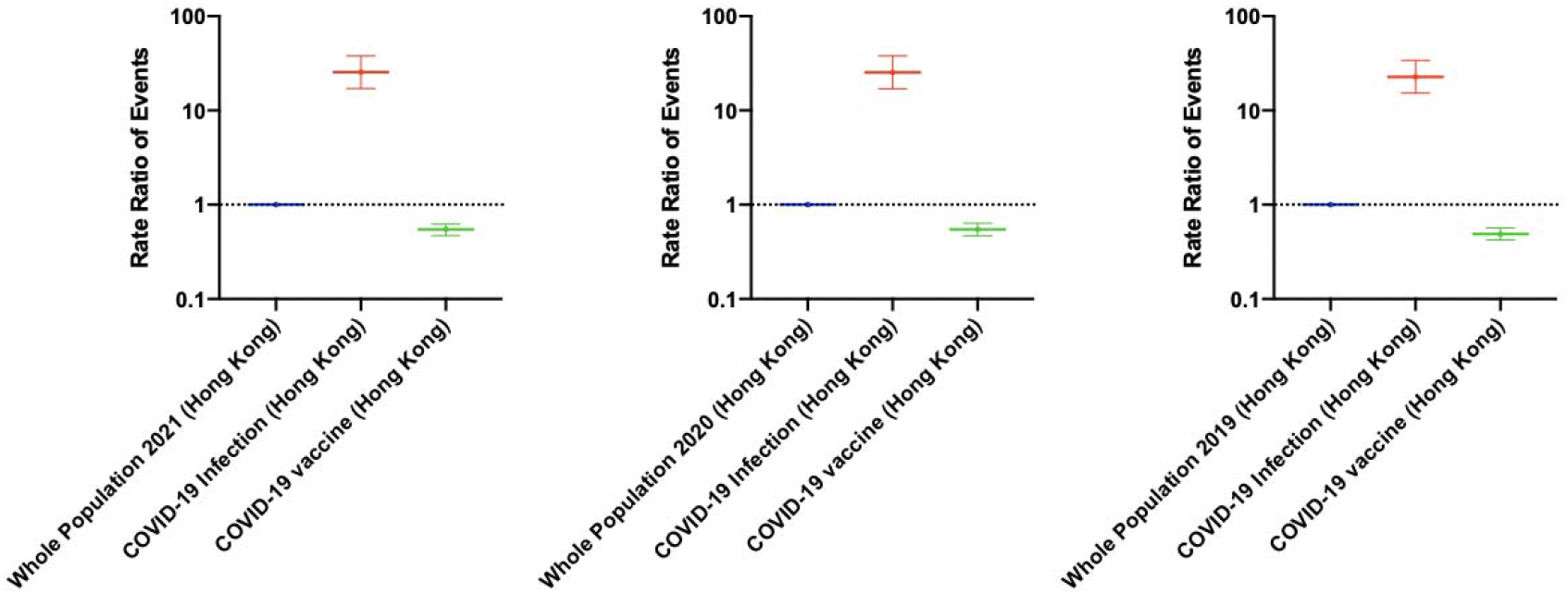
Sensitivity testing: The rate ratio of the acute myocardial infarction after COVID-19 infection and vaccination compared to the background rate across different years.

## Discussion

The present study found that patients infected with COVID-19 had a higher rate of AMI compared to those who were vaccinated. Furthermore, the rate of AMI among those who were vaccinated against COVID-19 was significantly lower than those who were not vaccinated. To the best of our knowledge, this study is the first study to demonstrate a lower rate of AMI in individuals vaccinated against COVID-19 in comparison to those who were infected with COVID-19.

### Comparison with previous studies

Although the literature on the relationship between AMI and COVID-19 vaccination is limited, with limited exploration of long-term follow-up data, a pooled analysis by Aye *et al*. revealed that the majority of AMI patients had received the Pfizer-BioNTech vaccine and developed symptoms after the first dose [15]. The patients generally had ischaemic ECG changes and raised cardiac troponin level [11]. Some studies have attributed cases of post-vaccine AMI to vaccine-associated immune thrombosis and thrombocytopenia/thrombosis [22, 23]. Furthermore, some vaccines, such as ChAdOx1, nCoV-19, and Ad26.COV2.S, demonstrated higher risks of AMI compared to other vaccines such as BNT162b2 [24]. However, it does not necessarily mean that COVID-19 vaccination is associated with higher risks of AMI. Indeed, similar proportion of individuals developed AMI in a randomised control trial of Moderna [14]. Meanwhile, no comparisons so far were made regarding the rate of AMI among the vaccinated individuals and those without vaccinations. We found that the rate of AMI among COVID-19 vaccinated individuals was significantly lower than in non-vaccinated individuals. This may be partly attributed to the difference in age between those who were vaccinated and those who were unvaccinated. Previously, it was reported that the mean age of the individuals vaccinated with CoronaVac (PiCoVacc) was 61.58 (SD: 11.08) and with BNT162b2 (Comirnaty) was 56.81 (SD: 13.43), which was a lot lower than the unvaccinated populations [25]. However, a population-based cohort study of 46 million adults found that rates of arterial or venous thrombotic events were generally lower after either vaccine compared with the unvaccinated, particularly in people aged ≥70 years [26].

By contrast, several studies have described relevant cardiac complications in COVID-19 patients with or without prior cardiovascular disease history [27-29]. COVID-19 was shown to significantly increase the risks of AMI [30, 31]. Notably, even the latter patients were more likely to experience AMI, heart failure, cardiogenic shock and life-threatening arrhythmias because of the SARS-CoV-2 infection’s direct impact on the cardiovascular system [32, 33]. Furthermore, as COVID-19 infections may cause high levels of inflammation, this could exacerbate the risks of blood clots in small blood vessels and long-term cardiovascular outcomes [34]. This is particularly concerning, as it was observed that during the COVID-19 outbreak, AMI was associated with eight times higher mortality [35]. In alignment with previous findings, compared to COVID-19 infection, the rate of AMI amongst the COVID-19 vaccinated population was significantly lower [30]. Furthermore, in Israel, compared to COVID-19 infection the rate of AMI immediately after COVID-19 vaccinated population was also significantly lower. Block *et al*. also demonstrated that COVID-19 patients were susceptible to greater risk of cardiac complications compared to after COVID-19 vaccination for both genders [36]. While this could be partly explained by the differences in the demographics between the three studied populations. The COVID-19 infected population was 69.11years old, which were higher than the 62.11 years old in the unvaccinated population and 59.40 years old in the vaccinated population previously reported [37]. Besides, it was also suggested that the COVID-19 infected patients were likely to have more cardiovascular disease history or cardiovascular risk factors compared to the general population [38]. Nevertheless, in the age-matched subgroup analysis, most of the COVID-19 patients under 70 years old who developed AMI did not have any remarkable cardiovascular disease history. Yet, the rate of AMI was still significantly higher amongst this subgroup. Our results further support the hypothesis that COVID-19 is an important risk factor for AMI in both patients with and without pre-existing cardiovascular disease [39, 40]. Furthermore, the COVID-19 vaccination may help protect against the COVID-19 caused AMI. This notwithstanding, the results of our previous study on the association of myopericarditis in vaccinated individuals also showed that the risks of adverse cardiovascular outcomes such as carditis were lower than those infected with SARS-CoV-2 [41] although the benefit of receiving two vaccine doses may not outweigh the risks of carditis for adolescent or male children [42].

### Potential underlying mechanisms

Although the precise mechanism has yet to be elucidated, several potential explanations exist regarding the association of COVID-19 with AMI. One such possibility pertains to the development of a pro-inflammatory state as a result of COVID-19 infection, which would result in the production of circulating cytokines and subsequent activation of inflammatory cells in atherosclerotic plaques. Resultantly, this increases plaque vulnerability, eventually leading to plaque rupture and coronary thrombosis [43, 44]. Indeed, the severity of COVID-19 pneumonia and increased D-dimer, C-reactive protein and lactate dehydrogenase levels was associated with increased severity of thromboembolic events [45]. Since COVID-19 infection results in the activation of the sympathetic system, tachycardia, hypotension, and hypoxemia in the setting of acute respiratory insufficiency, it might cause a mismatch between reduced oxygen supply and increased myocardial oxygen demand, leading to type 2 AMI [46]. Furthermore, it was suggested that COVID-19 infection would cause endothelial and microvascular injuries [47].

The majority of studies investigating AMI immediately after COVID-19 vaccination hypothesise a prothrombotic state as the underlying mechanism. While some claim that the vaccine causes an autoimmune response against platelets [48], others claim that AMI is caused by allergic vasospasms [49, 50]. This is exacerbated by the fact that elderly patients are frequently polymorbid, which may precipitate the onset of AMI [15]. Despite this, identifying this condition remains difficult because ischemic symptoms can be confused with mild reactions such as injection site soreness or chest pain [51]. Besides, the impacts of the vaccine on the COVID-19 vaccinated population in long term remained unknown.

### Clinical implications and the future

Our findings have significant clinical implications for vaccine acceptance in the general population and the safety profile of the COVID-19 vaccine at a population level. Prior to the pandemic, studies have demonstrated that influenza infection increases the risk of AMI, and the influenza vaccine effectively reduces the risk of AMI [52, 53]. Since then, the influenza vaccine has been proposed as a coronary intervention for preventing AMI which was likely to due to preventing influenza virus infection [54]. The COVID-19 vaccine may even have a similar effect. However, further studies are needed to demonstrate that the COVID-19 vaccine is protective against COVID-19-caused AMI.

By demonstrating that the rate of AMI was lower among those vaccinated with COVID-19, we add important evidence to the literature regarding the cardiovascular adverse effects of COVID-19 vaccines, which appears to be scarce at the moment. We emphasise the importance of suggesting vaccination to the public against COVID-19 because as it spreads across the globe, citizens will become more vulnerable to infection from SARS-CoV-2. This is concerning because, on a population level, the rate of AMI is much higher among those infected than among those vaccinated. These findings should raise clinicians’ awareness regarding the cardiac manifestations of COVID-19. Notably, the long-term effects of these acute cardiovascular manifestations of COVID-19 might become a healthcare challenge, resulting in increased mortality and morbidity [55]. Hence, strategies are needed to address the early prevention and diagnosis of at-risk COVID-19 patients.

### Strengths and limitations

Since the pandemic, the Hong Kong government has maintained a strict contact tracing policy in place [56]. As a result, our cohort likely represented the majority of infected patients in the general population [18, 57], allowing us to identify more post-COVID-19 AMI cases. However, there are certain limitations that should be acknowledged. Firstly, our cohort only included patients recruited from a single locality, so our findings are unable to account for geographical heterogeneity. Furthermore, because we only found a few studies that reported the rate of AMI post-vaccination, more cohort studies and randomised controlled trials are needed to determine the relationship between COVID-19 vaccination and AMI. Thirdly, the data reported by the Department of Health did not provide the number of AMI immediately post-vaccination, the number of AMI post-vaccination after the first and second dose of vaccination, the type of vaccination received, or the demographic landscape. Furthermore, the vaccinated population includes both COVID-19 infected and noninfected patients. As the COVID-19 infected patients account only for a small proportion amongst the vaccinated population, we believe this group can still be a fair comparison with the unvaccinated group. Vice versa, the COVID-19 infected patient may also be vaccinated. However, as the number of COVID-19 infected cases since the launch of the vaccination programme until the end of our study period was relatively small, most of the COVID-19 infected cases were not vaccinated. Lastly, as this was a retrospective study, the incidence of AMI after COVID-19 infection and vaccination can be underestimated compared to a prospective cohort study. More studies are needed to confirm the relationship.

## 5. Conclusion

At a population level, COVID-19 infection was associated with a significantly higher rate of AMI compared to the COVID-19 vaccinated population. The rate AMI after COVID-19 infection was also significantly higher than post COVID-19 vaccination across different countries. Further research is needed to examine the cardiovascular sequelae of COVID-19 infection and vaccination.

## Data Availability

All data produced in the present study are available upon reasonable request to the authors

## Supplementary Materials

The following are available online at www.mdpi.com/xxx/s1, Table S1: Supplementary Data

## Author Contributions

Conceptualization, Vassilios S Vassiliou, Bernard Man Yung Cheung and Gary Tse; Data curation, Oscar Hou-In Chou, Cheuk To Chung, Jiandong Zhou, Teddy Tai Loy Lee, Abraham Ka Chung Wai, Tong Liu and Gary Tse; Formal analysis, Oscar Hou-In Chou, Cheuk To Chung and Jiandong Zhou; Funding acquisition, Gary Tse; Investigation, Oscar Hou-In Chou and Danish Iltaf Satti; Methodology, Oscar Hou-In Chou, Teddy Tai Loy Lee and Gary Tse; Project administration, Tong Liu and Bernard Man Yung Cheung; Software, Oscar Hou-In Chou and Sharen Lee; Supervision, Tong Liu, Sharen Lee, Vassilios S Vassiliou, Bernard Man Yung Cheung and Gary Tse; Validation, Sharen Lee and Gary Tse; Writing – original draft, Oscar Hou-In Chou, Cheuk To Chung and Danish Iltaf Satti; Writing – review & editing, Oscar Hou-In Chou, Danish Iltaf Satti, Jiandong Zhou, Teddy Tai Loy Lee, Abraham Ka Chung Wai, Tong Liu, Sharen Lee, Vassilios S Vassiliou, Bernard Man Yung Cheung and Gary Tse.

## Institutional Review Board Statement

The study was conducted according to the guidelines of the Declaration of Helsinki, and ap-proved by the Institutional Review Board of the University of Hong Kong/Hospital Authority Hong Kong West Cluster (UW 20-250).

## Informed Consent Statement

Patient consent was waived due to the retrospective nature of the study and the data was deidentified.

## Data Availability Statement

Data available on request due to restrictions in privacy.

## Acknowledgments

Not applicable.

