## Supplementary Appendix for "Comparisons of the rate of acute myocardial infarction between COVID-19 patients and individuals received COVID-19 vaccines: a population-based study"

Supplementary Material

**Table S1.** Incidence rate and rate ratio of acute myocardial infarction after COVID-19 infection and vaccination in Israel.

| **Outcomes** | **COVID-19 infection**  **(Israel)[1]** | **COVID-19 vaccines**  **(Israel)[1]** |
| --- | --- | --- |
| Type of vaccines | BNT162b2 | BNT162b2 |
| Cases | 64 | 119 |
| Persons | 352400 | 1785570 |
| Follow-up period | 42 days | 42 days |
| Rate per million persons per 42 days (95% CI) | 182 (142, 232) | 66.6 (55.7, 79.7) |
| Rate ratio (95% CI) | 1 | 0.367 (0.271, 0.497) |

**Table S2.** Subgroup analysis 1. Incidence rate and rate ratio of AMI after COVID-19 infection and vaccination among elderlies across other countries.

| **Outcomes** | **COVID-19 infection**  **age >**=**70**  **(Israel)[1]** | **COVID-19 vaccines  age >= 70**  **(Israel)[1]** | **COVID-19 vaccines  age >= 75**  **(France)[2]** |
| --- | --- | --- | --- |
| Type of vaccines | BNT162b2 | BNT162b2 | BNT162b2 |
| Cases | 25 | 26 | 1277 |
| Persons | 11124 | 118728 | 7100000 |
| Follow-up period | 42 days | 42 days | 14 days |
| Rate per million persons per 42 days (95% CI) | 2247 (1523, 3316) | 219 (150, 320) | 540 (511, 570) |
| Rate ratio (95% CI) | 1 | 0.097 (0.067, 0.169) | 0.240 (0.162, 0.357) |

**Table S3.** Baseline characteristics for COVID AMI patients below the age of 70.

| **Demographic variables** | | **Mean ±  SD / Number and Percentage (n = 13)** | | |
| --- | --- | --- | --- | --- |
| **Age** | | 57.08 ± 16.56 | | |
| Male | | 10 | | 76.92 |
| **Comorbidities** |  | |  | |
| Hyperlipidaemia | | 0 | | 0 |
| Gastrointestinal bleeding | | 0 | | 0 |
| Acute myocardial infarction | | 1 | | 7.69 |
| VT/VF/SCD | | 0 | | 0 |
| Cognitive dysfunction | | 0 | | 0 |
| Anxiety disorder and depression | | 0 | | 0 |
| Ischemic heart disease | | 2 | | 15.38 |
| Sleep disorders | | 0 | | 0 |
| Hip fractures | | 0 | | 0 |
| Accident fall | | 1 | | 7.69 |
| Alcoholism | | 0 | | 0 |
| Tobacco use and dependence | | 0 | | 0 |
| Obesity | | 0 | | 0 |

**Table S4.** Subgroup analysis 2. The rate ratio of the acute myocardial infarction events after COVID-19 infection under 70 years old and COVID-19 vaccination in Hong Kong.

| **Outcomes** | **Non COVID-19 vaccine**  **(Hong Kong, China)[3]** | **Covid-19 infection**  **age <= 70**  **(Hong Kong, China)** | **COVID-19 vaccine**  **(Hong Kong, China)[3]** |
| --- | --- | --- | --- |
| Type of vaccines | - | - | CoronaVac  BNT162b2 |
| Cases | 405 | 13 | 231 |
| Persons | 2500000 | 10276 | 4914894 |
| Follow-up period | 28 days | 28 days | 28 days |
| Rate per million persons or doses per 28 days (95% CI) | 162 (147, 179) | 1265 (749, 2163) | 47.0 (41.3, 53.5) |
| Rate ratio (95% CI) | 1 | 7.81 (4.50, 13.6) | 0.290 (0.247, 0.341) |

**Table S5.** Sensitivity testing: The rate ratio of the AMI events after COVID-19 infection and vaccination compared to the background rate across different years.

| **Outcomes** | **Whole population 2021**  **(Hong Kong, China)[3]** | **Covid-19 infection**  **(Hong Kong, China)** | **COVID-19 vaccine**  **(Hong Kong, China)[3]** |
| --- | --- | --- | --- |
| Type of vaccines | - | - | CoronaVac  BNT162b2 |
| Cases | 636 | 25 | 231 |
| Persons | 7414894 | 11441 | 4914894 |
| Follow-up period | 28 days | 28 days | 28 days |
| Rate per million persons or doses per 28 days (95% CI) | 85.7 (79.4, 92.7) | 2884  (2055, 4048) | 47.0 (41.3, 53.5) |
| Rate ratio (95% CI) | 1 | 25.5  (17.1, 38.0) | 0.55  (0.47, 0.62) |
| **Outcomes** | **Whole population 2020**  **(Hong Kong, China)[3]** | **Covid-19 infection**  **(Hong Kong, China)** | **COVID-19 vaccine**  **(Hong Kong, China)[3]** |
| Type of vaccines | - | - | CoronaVac  BNT162b2 |
| Cases | 647 | 25 | 231 |
| Persons | 7523256 | 11441 | 4914894 |
| Follow-up period | 28 days | 28 days | 28 days |
| Rate per million persons or doses per 28 days (95% CI) | 86.0  (79.6, 92.9) | 2884  (2055, 4048) | 47.0 (41.3, 53.5) |
| Rate ratio (95% CI) | 1 | 25.4  (17.0, 37.9) | 0.55  (0.47, 0.64) |
| **Outcomes** | **Whole population 2019**  **(Hong Kong, China)[3]** | **Covid-19 infection**  **(Hong Kong, China)** | **COVID-19 vaccine**  **(Hong Kong, China)[3]** |
| Type of vaccines | - | - | CoronaVac  BNT162b2 |
| Cases | 724 | 25 | 231 |
| Persons | 7541667 | 11441 | 4914894 |
| Follow-up period | 28 days | 28 days | 28 days |
| Rate per million persons or doses per 28 days (95% CI) | 96.0  (89.3, 103) | 2884  (2055, 4048) | 47.0 (41.3, 53.5) |
| Rate ratio (95% CI) | 1 | 22.8  (15.3, 33.9) | 0.49  (0.42, 0.57) |

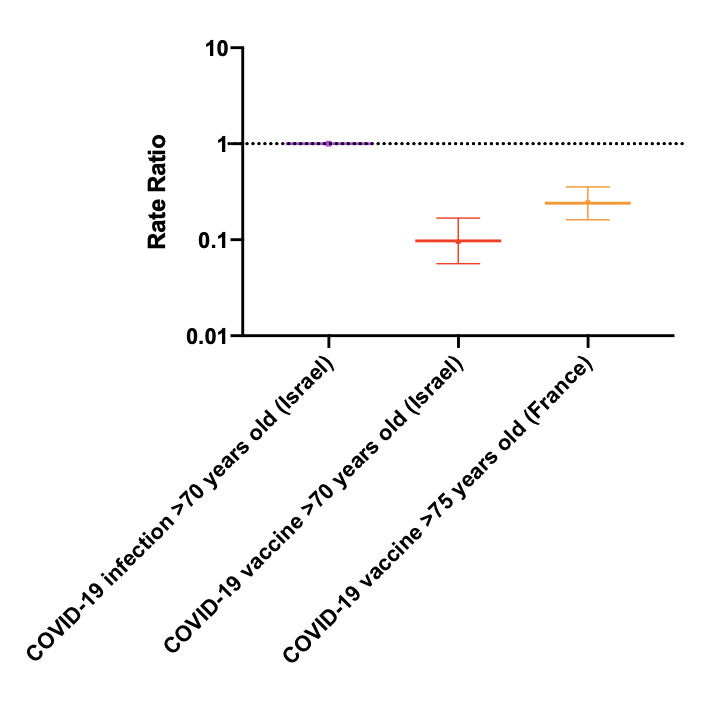

**Supplementary Figure 1. Subgroup analysis 1. Incidence rate and rate ratio of AMI after COVID-19 infection and vaccination among elderlies across other countries.**
